# Effect of Machine Learning on Anaesthesiology Clinician Prediction of Postoperative Complications: The Perioperative ORACLE Randomised Clinical Trial

**DOI:** 10.1101/2024.05.22.24307754

**Authors:** Bradley A Fritz, Christopher R King, Mohamed Abdelhack, Yixin Chen, Alex Kronzer, Joanna Abraham, Sandhya Tripathi, Arbi Ben Abdallah, Thomas Kannampallil, Thaddeus P Budelier, Daniel Helsten, Arianna Montes de Oca, Divya Mehta, Pratyush Sontha, Omokhaye Higo, Paul Kerby, Stephen H. Gregory, Troy S. Wildes, Michael S Avidan

## Abstract

**Background:** Anaesthesiology clinicians can implement risk mitigation strategies if they know which patients are at greatest risk for postoperative complications. Although machine learning models predicting complications exist, their impact on clinician risk assessment is unknown.

**Methods:** This single-centre randomised clinical trial enrolled patients age ≥18 undergoing surgery with anaesthesiology services. Anaesthesiology clinicians providing remote intraoperative telemedicine support reviewed electronic health records with (assisted group) or without (unassisted group) also reviewing machine learning predictions. Clinicians predicted the likelihood of postoperative 30-day all-cause mortality and postoperative acute kidney injury within 7 days. Area under the receiver operating characteristic curve (AUROC) for the clinician predictions was determined.

**Results:** Among 5,071 patient cases reviewed by 89 clinicians, the observed incidence was 2% for postoperative death and 11% for acute kidney injury. Clinician predictions agreed with the models more strongly in the assisted versus unassisted group (weighted kappa 0.75 versus 0.62 for death [difference 0.13, 95%CI 0.10-0.17] and 0.79 versus 0.54 for kidney injury [difference 0.25, 95%CI 0.21-0.29]). Clinicians predicted death with AUROC of 0.793 in the assisted group and 0.780 in the unassisted group (difference 0.013, 95%CI −0.070 to 0.097). Clinicians predicted kidney injury with AUROC of 0.734 in the assisted group and 0.688 in the unassisted group (difference 0.046, 95%CI −0.003 to 0.091).

**Conclusions:** Although there was evidence that the models influenced clinician predictions, clinician performance was not statistically significantly different with and without machine learning assistance. Further work is needed to clarify the role of machine learning in real-time perioperative risk stratification.

**Trial Registration:** ClinicalTrials.gov NCT05042804

## INTRODUCTION

Postoperative mortality remains a major problem worldwide, with more than four million people dying within 30 days after surgery annually.^1^ Many deaths are preceded by complications such as acute kidney injury (AKI), respiratory failure, or adverse cardiac events.^2–4^ Early recognition of these postoperative risks can facilitate changes in intraoperative or postoperative management to prevent complications or detect them sooner.^5,6^ Although data are available to identify patients at greatest risk for postoperative complications, accurate real-time risk assessment is difficult. First, the volume of data available during surgery often exceeds human information processing capacity, especially considering that the anaesthesiology clinician must concurrently perform multiple clinical care tasks.^7–9^ Second, anaesthesiology clinicians frequently succumb to biases and other cognitive errors when synthesizing available data to make clinical decisions.^10^ These difficulties with risk assessment likely increase for attending anaesthesiologists who oversee the care of multiple surgical patients simultaneously.

Researchers have proposed using machine learning (ML) models to mitigate the known limitations in anaesthesiology clinician risk assessment. Currently, ML models are available that predict postoperative mortality,^11–15^ AKI,^16–18^ and other postoperative complications^19,20^ with moderate-to-high discrimination. However, it has not been ascertained whether anaesthesiology clinicians would incorporate such models into their clinical practice to identify more accurately which patients are at risk for complications and therefore might benefit from risk mitigation strategies or enhanced monitoring. The objectives of this study were to determine whether anaesthesiology clinicians (i) incorporate information from ML predictions into their determination of postoperative patient risk, and (ii) can predict postoperative complications more accurately with access to ML model support than without ML model support.

## METHODS

### Overall Design

We conducted the Perioperative Outcome Risk Assessment with Computer Learning Enhancement (Perioperative ORACLE) single-centre randomized trial (ClinicalTrials.gov NCT05042804, registered 9/13/2021). This trial was nested within the TECTONICS (Telemedicine Control Tower for the Operating Room: Navigating Information, Care and Safety) randomized clinical trial (NCT03923699). TECTONICS tested the effect of an anaesthesiologist-staffed remote intraoperative telemedicine intervention (the Anesthesiology Control Tower, or ACT) on postoperative 30-day mortality, delirium, respiratory failure, and AKI. The ACT intervention consisted of reactive support in response to rule-based physiologic alerts and proactive support via comprehensive patient case reviews with communication of risk assessment and treatment recommendations to the operating room anaesthesiology team. For patients co-enrolled in Perioperative ORACLE, the comprehensive patient case review was randomized 1:1 to be performed with or without access to ML models predicting postoperative complications. The institutional review board at Washington University School of Medicine approved both TECTONICS (approval #201903026) and Perioperative ORACLE (#202108022) with a waiver of informed consent. Protocols for both trials have been published.^21,22^ This manuscript is written in accordance with the CONSORT-AI guidelines.^23^

### Setting and Participants

This trial was conducted at Barnes-Jewish Hospital, a university-affiliated, tertiary care hospital in Saint Louis, Missouri. Patients were included in Perioperative ORACLE if they were enrolled in TECTONICS and had a comprehensive patient case review performed between 9/13/2021 and 9/30/2022. The selection of cases to review was at the discretion of the clinical staff in the ACT. Inclusion criteria for TECTONICS included age ≥ 18, surgery with anaesthesiology services at Barnes-Jewish Hospital, and surgery starting 7am-4pm Monday-Friday. ACT clinicians were members of the TECTONICS research team and included attending anaesthesiologists, resident physicians, and nurse anaesthetists.

### ML Model Intervention

The intervention included ML models predicting death from any cause within 30 days after surgery and postoperative AKI defined using KDIGO creatinine criteria.^24^ Input features were selected by clinical experts and included demographic characteristics, comorbid conditions, preoperative vital signs, surgical service, functional capacity, and laboratory results. Models of varying architectures were trained using a retrospective cohort of surgical patients from the same institution, and the model with largest area under receiver operating characteristic curve (AUROC) in a randomly held-out validation set was selected. At the start of the trial, we used random forest models trained using approximately 110,000 patients who underwent surgery 2012-2016 (model version 1). Because of a change in the electronic health record vendor in 2018 leading to shifts in several data elements, new models were trained using 84,000 patients who underwent surgery 2018-2020, adding the planned surgical procedure (in addition to surgical service) as an input feature. After retraining, the best performing model was a gradient boosted decision tree (model version 2). Model version 2 was deployed with an updated interface in February 2022. No further changes to model architecture or recalibrations were performed during the trial.

Models were implemented in Python. Computations were performed on a HIPAA-compliant, university-maintained server with a near-live data feed from the electronic health record. Missing values were imputed with the population median for continuous variables or treated as a “missing” category for categorical variables. Model outputs were displayed to ACT clinicians on a secure, password-protected web application (Supplement Figure 1). Information displayed included the predicted risk of the complication (0%-100%), graphs showing change in predicted risk over time, and feature contribution estimates obtained using Shapley values. The design of the display was informed by a needs assessment study.^25^

### Procedures

As part of their role in the parent TECTONICS trial, ACT clinicians conducted patient case reviews intraoperatively, generally within the first 60 minutes. The clinician reviewed pertinent data in the electronic health record, such as preoperative notes, laboratory results, diagnostic studies, and the start of the current anaesthesia record. The clinician completed an electronic case review form within a customized version of AlertWatch:OR software (BioIntelliSense, Golden, CO). The case review form included (among other elements) a rating of how likely the patient was to experience each complication on a five-point Likert scale: very low risk, low risk, average risk, high risk, or very high risk.

### Randomization

Case reviews were randomized 1:1 without restriction to be performed with or without utilizing the ML model output. The allocation was displayed when the ACT clinician opened the case review form in AlertWatch. The allocation sequence was a pseudo-random function of a back-end surgical encounter identifier. If randomized to the ML-assisted group, clinicians reviewed the ML web application before completing the case review form. If randomized to the ML-unassisted group, clinicians completed the case review form immediately. Clinicians could not be blinded to allocation.

To assess compliance with randomization, clinicians self-reported if they reviewed the ML model output during the case review. If the clinicians reviewed the ML model output, then they were also asked whether they found the ML predictions surprising and whether they agreed or disagreed with the ML predictions.

### Outcomes

The co-primary outcomes were accuracy of clinician predictions of postoperative 30-day all-cause mortality and postoperative AKI. True complication status was defined using electronic health record queries. 30-day all-cause mortality was defined using vital status as documented in the electronic health record. AKI was defined as an increase in creatinine ≥0.3 mg/dL within 48 hours or ≥1.5 times baseline within 7 days.^24^ If creatinine was not checked preoperatively, then the upper limit of the laboratory’s reference range was used as the baseline value. If creatinine was not checked postoperatively, then AKI was assumed to be absent. AKI was not defined for patients with a baseline creatinine >4.0 mg/dL, patients meeting the creatinine criteria for AKI preoperatively, patients receiving dialysis preoperatively, or patients undergoing a dialysis access procedure or kidney transplant.

### Statistical Methods and Sample Size

All analyses were conducted using R version 4.2.3.^26^ Descriptive statistics included frequencies (percentage) for categorical variables and either mean (standard deviation) or median (interquartile range) for continuous variables depending on the distribution. Agreement between clinician predictions and ML predictions was quantified using weighted kappa, with quadratic weights. For each co-primary outcome, two logistic regression models were constructed: one using case reviews in the ML-assisted group and another using case reviews in the ML-unassisted group. Each logistic regression used clinician predictions (a 5-level categorical variable) as the independent variable and true complication status as the dependent variable. DeLong’s test was used to compare the AUROC of the model using ML-assisted cases to the AUROC of the model using ML-unassisted cases.^27^ The sample size calculation was previously published.^22^

The primary analysis followed the intention-to-treat principle and included all patients for whom clinician predictions were available and the true complication status was known. Secondary per-protocol and as-treated analyses were also performed. For AKI, a sensitivity analysis excluded patients with no postoperative creatinine measurement. As exploratory analyses, the primary analysis was repeated in subgroups defined by sex, race, ML model version (1 or 2), clinician background, and randomization allocation from the parent TECTONICS trial. An additional post-hoc subgroup was patients where the ML model correctly predicted the complication status (when the model prediction 60 minutes after start of surgery was dichotomized at the value that maximized the Youden index).

## RESULTS

Between 9/13/2021 and 9/30/2022, 5,071 patient cases were included in the trial, with 2,536 case reviews randomized to the ML-assisted group and 2,535 to the ML-unassisted group (Figure 1). Baseline characteristics of the randomized patients are shown in Table 1. In total, 89 distinct anaesthesiology clinicians participated. The trial ended because the target enrolment was achieved.

**Figure 1.**
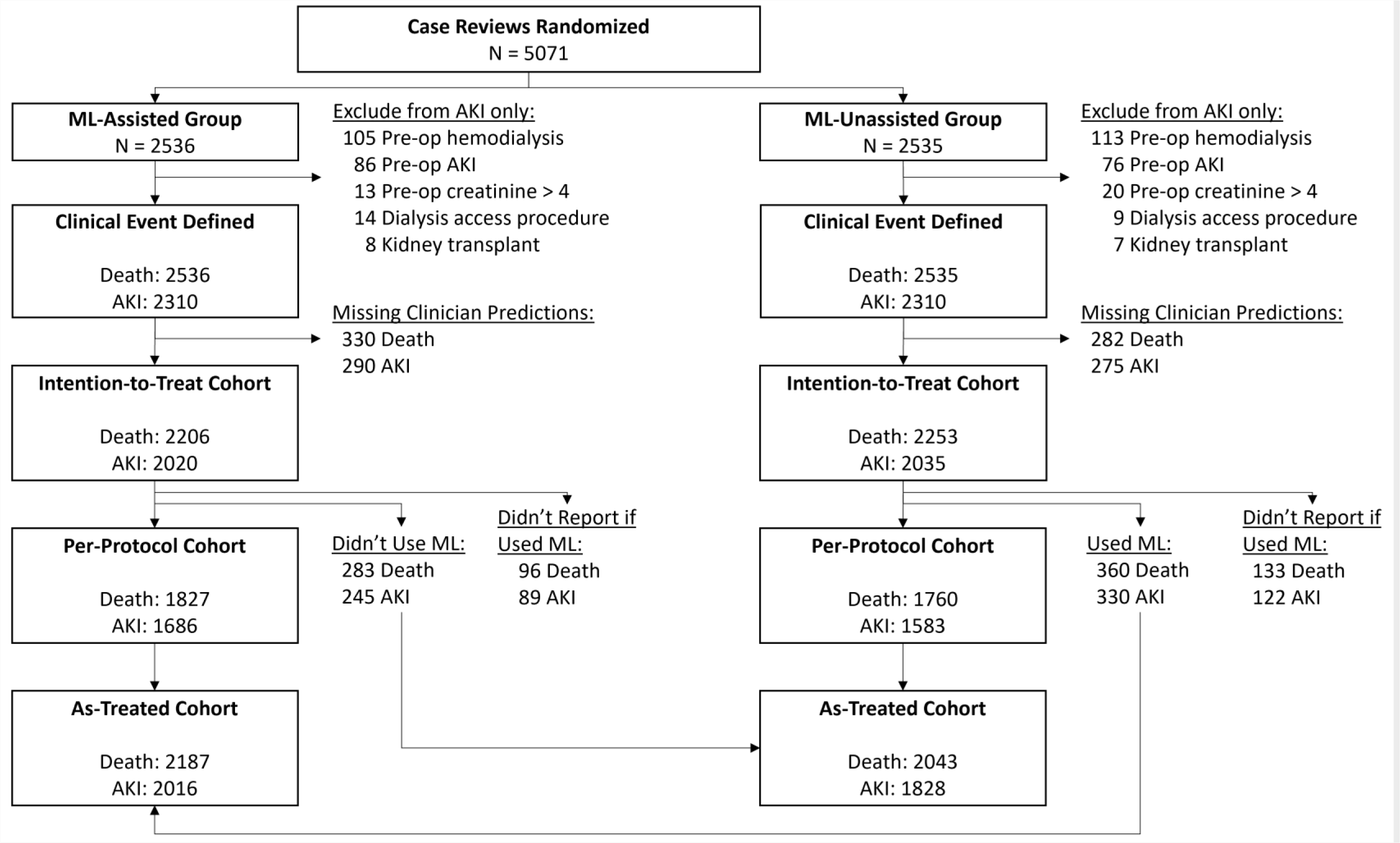
CONSORT Flow Diagram. Abbreviations: AKI = acute kidney injury. ML = machine learning.

**Table 1.** Patient and Clinician Characteristics for Randomized Case Reviews.

| **Variable** | **ML-Assisted Group**<br>**(N = 2536)** | **ML-Unassisted Group**<br>**(N = 2535)** |
| --- | --- | --- |
| Age, mean (standard deviation) | 58.2 (15.7) | 57.8 (16.2) |
| Sex |  |  |
| Female | 1315 (52%) | 1338 (53%) |
| Male | 1221 (48%) | 1197 (47%) |
| Race |  |  |
| Black or African American | 543 (21%) | 507 (20%) |
| White | 1925 (76%) | 1942 (77%) |
| Other | 44 (2%) | 55 (2%) |
| Unknown | 24 (1%) | 31 (1%) |
| Ethnicity |  |  |
| Hispanic | 48 (2%) | 43 (2%) |
| Non-Hispanic | 2455 (97%) | 2461 (97%) |
| Unknown | 33 (1%) | 31 (1%) |
| Surgery Service |  |  |
| General Surgery | 612 (24%) | 602 (24%) |
| Orthopedics | 408 (16%) | 421 (17%) |
| Obstetrics/Gynecology | 291 (11%) | 288 (11%) |
| Neurosurgery | 241 (10%) | 267 (11%) |
| Urology | 232 (9%) | 231 (9%) |
| Cardiothoracic | 212 (8%) | 220 (9%) |
| Vascular | 211 (8%) | 182 (7%) |
| Otolaryngology | 188 (7%) | 200 (8%) |
| Plastics | 95 (4%) | 92 (4%) |
| Other | 44 (2%) | 29 (1%) |
| ASA Physical Status |  |  |
| 1 | 65 (3%) | 65 (3%) |
| 2 | 764 (30%) | 760 (30%) |
| 3 | 1370 (54%) | 1411 (56%) |
| 4 | 316 (13%) | 279 (11%) |
| 5 | 10 (< 1%) | 13 (1%) |
| ASA Emergency Status |  |  |
| Not Emergency | 2452 (97%) | 2471 (98%) |
| Emergency | 73 (3%) | 57 (2%) |
| TECTONICS Randomization |  |  |
| Telemedicine Intervention | 1276 (50%) | 1236 (49%) |
| Usual Care | 1260 (50%) | 1299 (51%) |
| Clinical Experience of ACT Clinician |  |  |
| MD/DO - Attending | 766 (30%) | 779 (31%) |
| MD/DO - CA3 Resident | 666 (26%) | 670 (26%) |
| MD/DO - Intern Level | 682 (27%) | 648 (26%) |
| CRNA | 292 (12%) | 290 (11%) |
| SRNA | 130 (5%) | 148 (6%) |

Abbreviations: ML = Machine learning. ASA = American Society of Anesthesiologists. TECTONICS = Telemedicine Control Tower for the Operating Room: Navigating Information, Care and Safety. ACT = Anesthesiology Control Tower. CRNA = Certified registered nurse anaesthetist. SRNA = Student registered nurse anaesthetist.

### Prospective Performance of ML Models

Overall, the ML model predicting death performed with AUROC 0.807 (95%CI 0.768-0.847), whereas the model predicting AKI performed with AUROC 0.766 (95%CI 0.746-0.787). Both models exhibited variation in performance month-to-month but showed improvement after the new model versions were implemented in February 2022 (Supplement Figure 2). Prospective performance of all models was reduced compared to performance in the retrospective holdout validation datasets (Supplement Table 1).

### Patterns of Clinician Predictions

Clinician predictions were more likely to match the ML predictions (i.e., fall into the same risk category on the 5-point Likert scale) in the ML-assisted group compared to the ML-unassisted group when predicting death (weighted kappa 0.75 versus 0.62, difference 0.13 [95%CI 0.10-0.17], Figure 2) and AKI (weighted kappa 0.79 versus 0.54, difference 0.25 [95%CI 0.21-0.29], Figure 3). These patterns were seen regardless of whether the ML prediction was correct or incorrect when dichotomized at the value that maximized the Youden index (Supplement Figures 3-6, Supplement Tables 2-3). In the ML-assisted group, clinician and ML predictions for both outcomes were most likely to match when clinicians self-reported not being surprised by the ML (Supplement Figures 7-8, Supplement Tables 4-5).

**Figure 2.**
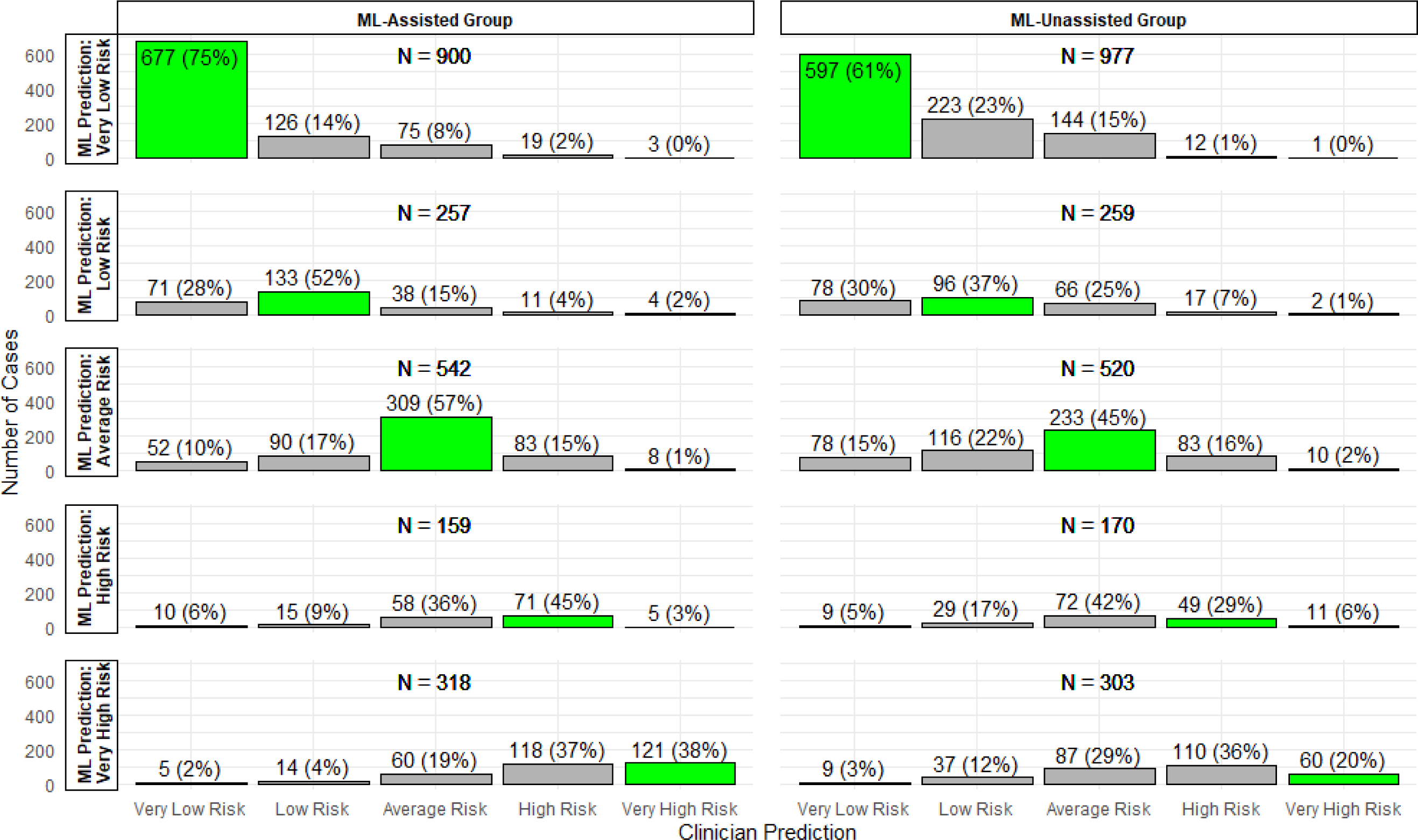
Distribution of Clinician Predictions for Postoperative Death. Stratified by treatment allocation (ML-unassisted group versus ML-assisted group) and by ML prediction. Green bars represent cases where the clinician prediction matched the categorical ML prediction.

**Figure 3.**
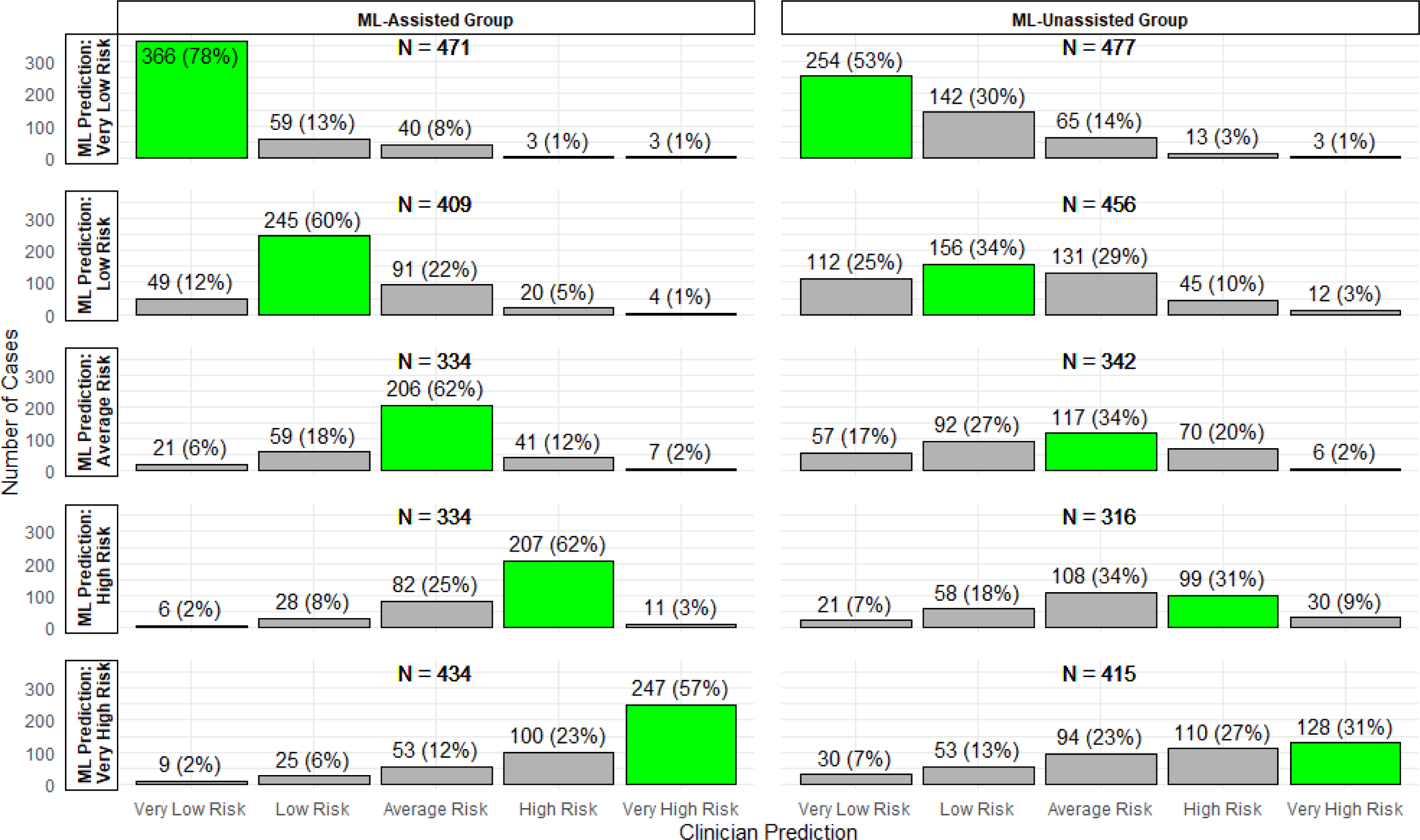
Distribution of Clinician Predictions for Postoperative Acute Kidney Injury. Stratified by treatment allocation (ML-unassisted group versus ML-assisted group) and by ML prediction. Green bars represent cases where the clinician prediction matched the categorical ML prediction.

**Figure 4.**
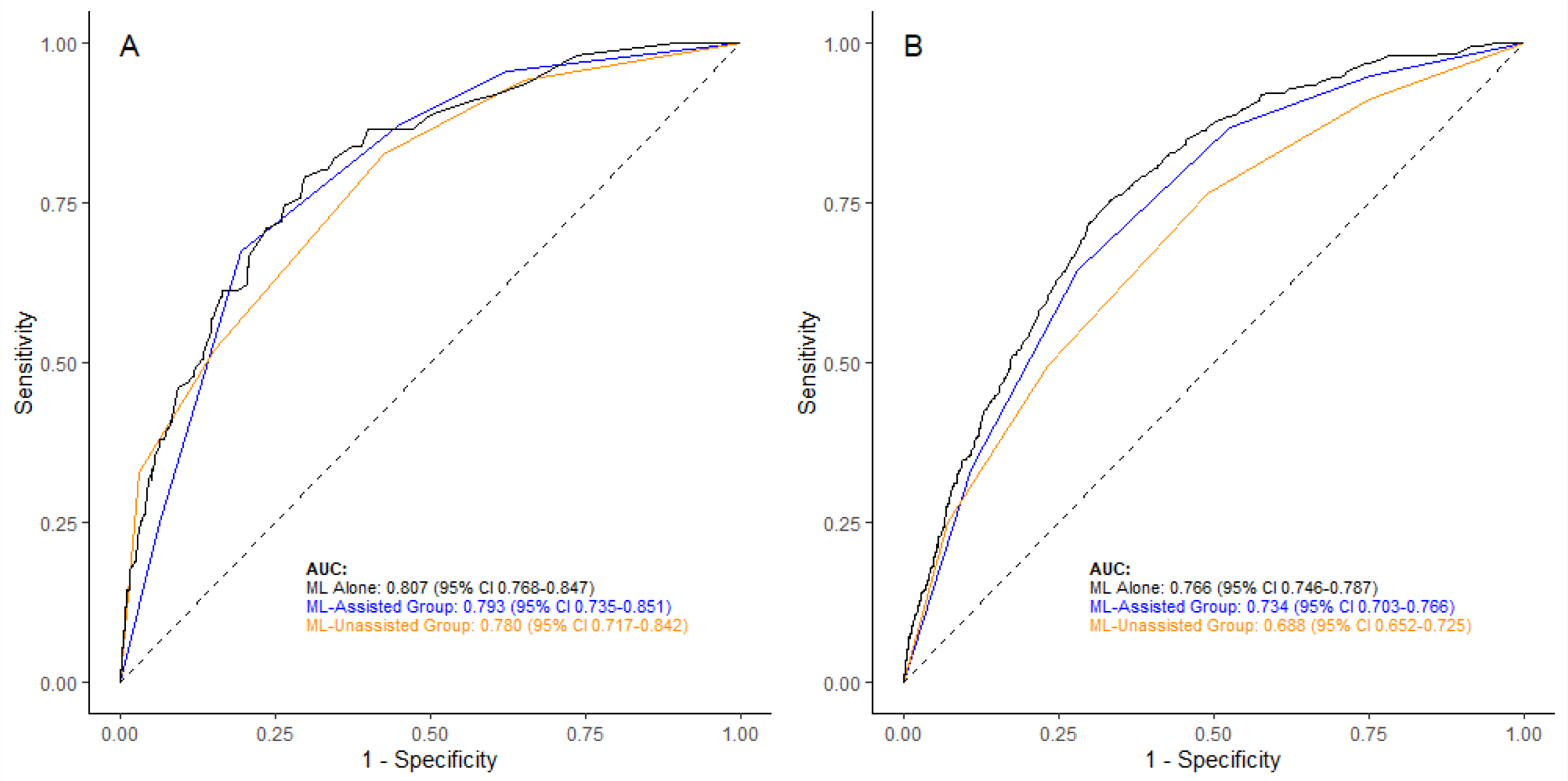
Receiver Operating Characteristic Curves. (A) Prediction of postoperative death within 30 days. (B) Prediction of postoperative acute kidney injury. Abbreviations: AUC = Area under curve. ML = Machine learning

### Performance for Prediction of Death

The primary analysis for death included 4,459 patients, of whom 98 (2.2%) died within 30 days after surgery. Clinicians in the ML-assisted group predicted mortality with AUROC 0.793 (95%CI 0.735-0.851), while clinicians in the ML-unassisted group predicted mortality with AUROC 0.780 (95%CI 0.717-0.842, Figure 2A). The difference in AUROC between the two groups was 0.013 (95%CI −0.070 to 0.097, *p*=0.76). The positive predictive value of a “high risk” or “very high risk” prediction was 6.9% in the ML-assisted group and 7.5% in the ML-unassisted group (Supplement Table 6). There was no difference in AUROC between groups in the per-protocol, as-treated, and pre-specified subgroup analyses (Supplement Figure 9). However, the AUROC was higher in the ML-assisted group than in the ML-unassisted group for patients where the dichotomized ML prediction was correct.

### Performance for Prediction of AKI

The primary analysis for AKI included 4,055 patients, of whom 450 (11.1%) experienced AKI. Clinicians in the ML-assisted group predicted AKI with AUROC 0.734 (95%CI 0.703-0.766), while clinicians in the ML-unassisted group predicted AKI with AUROC 0.688 (95%CI 0.652-0.725, Figure 2B). The difference in AUROC between the two groups was 0.046 (95%CI −0.003 to 0.091, *p*=0.06). The positive predictive value of a “high risk” or “very high risk” prediction was 23.1% in the ML-assisted group and 20.2% in the ML-unassisted group (Supplement Table 7). There was no difference in AUROC between the ML-assisted group and the ML-unassisted group in the per-protocol, as-treated, and pre-specified subgroup analyses (Supplement Figure 10). However, the AUROC was higher in the ML-assisted group than in the ML-unassisted group for patients where the dichotomized ML prediction was correct and lower when the dichotomized ML prediction was incorrect.

## DISCUSSION

In this single-centre, randomized clinical trial, anaesthesiology clinicians predicted postoperative death within 30 days with high discrimination both with and without access to ML predictive algorithms. The clinicians predicted postoperative AKI with moderate discrimination, and the difference in discrimination between the ML-assisted group and the ML-unassisted group was not statistically significant. For both models, prospective model performance was reduced compared to performance during validation in a retrospective cohort.

A few previous studies have examined ML and clinician performance in perioperative prediction tasks. In a simulation study, 20 intensivists predicted 2 out of 6 postoperative complications more accurately for 150 patients after reviewing the *MySurgeryRisk* tool than before reviewing it.^28^ However, there was no change in prediction accuracy by surgeons during live deployment of this risk tool.^29^ In another simulation study, 5 anaesthesiologists predicted intraoperative hypoxemia with greater discrimination using an ML model than not using it.^30^ Finally, the 68-patient HYPE trial reported reduced time-weighted average hypotension during elective non-cardiac surgery using an early warning system than without it,^31^ although no difference was found in subsequent evaluation by different investigators.^32^ These trials did not explicitly measure clinician predictions, but the observed effects were presumably mediated by changes in anaesthesiologist anticipation of hypotension.

There are key differences between our trial and these previous trials. In the simulation studies, clinicians had no other duties besides prediction, whereas in ORACLE clinicians simultaneously provided telemedicine support for ongoing surgeries. In some studies that used historical cases, the outcome incidence was artificially increased by up-sampling positive cases, which can impact clinician performance. Some trials included few clinicians, who may have developed expertise in using the ML model output over time, whereas ORACLE included many anaesthesiology clinicians with different backgrounds and levels of experience. Taken as a whole, the literature surrounding perioperative outcome prediction resembles the literature elsewhere in medicine, where about 50% of studies report improvements in diagnostic performance by ML-assisted clinicians compared to clinicians alone.^33^

Importantly, ORACLE clinician predictions in the ML-assisted group were more likely to match the ML model output compared to the ML-unassisted group. This implies that ML impacted the clinicians’ predictions. However, this impact occurred equally regardless of whether the ML was correct or incorrect (Supplement Tables 2-3), raising concern for “automation bias” where ML output is quickly accepted rather than critically reviewed. Therefore, the lack of statistically significant differences in discrimination between groups may be driven at least partly by poorer ML model performance during prospective deployment compared to retrospective evaluation. Reasons for the decrement in performance might include data drift or concept drift. Data drift refers to changes in the distribution of input features over time, either due to true changes in the patient population (e.g., increasing age and comorbidity burden of surgical patients over time^34^) or changes in data capture (e.g., new data field or increased missing rates). Concept drift refers to changes in the target variable being predicted (e.g., distribution of causes of death changing over time). Both factors can change the relationships between input features and the prediction target, causing degradation of model performance. Poorer prospective performance can also be a sign of unintentional overfitting due to multiple experiments during algorithm development.

This trial has numerous strengths. First, nesting within the TECTONICS trial allowed many patient cases to be reviewed efficiently. Second, patient cases were reviewed by many anaesthesiology clinicians with differing backgrounds, including attending anaesthesiologists, resident physicians, and nurse anaesthetists. Third, cases were reviewed in real-time during surgery with ML models utilizing live data feeds from the electronic health record. Using real-time case reviews rather than simulations means the trial more closely captured how ML models might be used at the bedside. Fourth, the ML model user interface was designed based on input obtained from the anaesthesiology clinicians who would be using it.^25^ Fifth, the ML model performed on-par or better than expert clinicians despite having access to many fewer input features. This points to a role for ML in data review, especially in teams where some members have less experience or different content area expertise.

This trial also has limitations. First, case reviews were conducted in the context of a telemedicine intervention rather than at the patient’s bedside. However, clinicians reported that the workflow they used for evaluating patients in the ACT closely mimics the workflow they use before taking patients to the operating room for in-person anaesthesiology care.^25^ Second, we were unable to measure how much time clinicians spent reviewing each case. ML models may provide value if they allow clinicians to arrive at the same risk assessment more quickly, even if accuracy remains unchanged. Third, clinicians assessed risk using a five-point Likert scale rather than a continuous scale, which may have decreased our ability to detect differences in discrimination between the groups. However, we believe that the 5-point scale captures the most clinically relevant risk assessment. Fourth, this work was conducted at a single academic medical centre. The results may be different in other contexts. Fifth, discrimination was somewhat lower in the ML-assisted group compared to the ML models alone. This may indicate insufficient trust in the ML models. Additional work is needed to understand reasons for lack of clinician trust, but possible explanations might include inadequate explanations of contributing features or poor workflow integration. Sixth, our results speak only to differences in discrimination using ML, not differences in calibration, which is also important for clinical decision making. Seventh, AKI was assumed to be absent if creatinine was not measured postoperatively. However, results were similar in a sensitivity analysis excluding these patients. Eighth, although our study is large, the confidence intervals on differences in discrimination are relatively wide, especially for death, where the number of events is smaller.

In conclusion, this trial demonstrated that anaesthesiology clinicians predicted postoperative death and AKI without statistically significant difference in discrimination whether or not they had access to ML predictive algorithms. The ML models appeared to impact clinician predictions, improving clinician discrimination when the ML was correct but reducing clinician discrimination when the ML was incorrect. Although caution is needed to prevent poorly performing models from leading clinicians astray, this work suggests there may be a role for well-calibrated models to improve clinician risk assessment.

## Supporting information

Supplementary Digital Content

CONSORT-AI Checklist

## Data Sharing Statement

The Washington University Human Research Protection Office did not permit sharing of individual patient-level data due to enrollment with a waiver of informed consent. However, group cross-tabulations with sufficient detail to replicate the analyses in this manuscript are available on request.

## Author Contributions

BAF: Study conception and design, acquisition of data, data analysis and interpretation, drafting and critical revision of manuscript.

CRK: Study conception and design, acquisition of data, data analysis and interpretation, critical revision of manuscript.

MA: Design and implementation of machine learning models, data interpretation, critical revision of manuscript.

YC: Study conception and design, data interpretation, critical revision of manuscript.

AK: Acquisition of data, data interpretation, critical revision of manuscript.

JA: Study conception and design, data interpretation, critical revision of manuscript.

ST: Design and implementation of machine learning models, data interpretation, critical revision of manuscript.

ABA: Study design, data interpretation, critical revision of manuscript.

TK: Study conception and design, data interpretation, critical revision of manuscript.

TPB: Acquisition of data, data interpretation, critical revision of manuscript.

DH: Study conception and design, data interpretation, critical revision of manuscript.

AMdO: Acquisition of data, data interpretation, critical revision of manuscript.

DM: Acquisition of data, data interpretation, critical revision of manuscript.

PS: Acquisition of data, data interpretation, critical revision of manuscript.

OH: Study design, data interpretation, critical revision of manuscript.

PK: Study design, data interpretation, critical revision of manuscript.

SHG: Study design, data interpretation, critical revision of manuscript.

TSW: Study conception and design, data interpretation, critical revision of manuscript.

MSA: Study conception and design, acquisition of data, data interpretation, critical revision of manuscript.

