## Supplementary Digital Content for "Effect of Machine Learning on Anaesthesiology Clinician Prediction of Postoperative Complications: The Perioperative ORACLE Randomised Clinical Trial"

**Supplementary Digital Content for "The Effect of Machine Learning Decision Support on Intraoperative Anesthesiology Clinician Prediction of Postoperative Complications: The Perioperative ORACLE Randomized Clinical Trial"**

Supplement Figure 1. Screenshot of User Interface

Supplement Figure 2. Prospective Performance of Machine Learning Models Over Time

Supplement Table 1. Retrospective and Prospective Performance of Machine Learning Models

Supplement Figure 3. Clinician Predictions for Postoperative Death in Subgroup where Dichotomized ML Prediction Was Correct

Supplement Figure 4. Clinician Predictions for Postoperative Death in Subgroup where Dichotomized ML Prediction Was Incorrect

Supplement Table 2. Agreement Between Clinician and ML Predictions of Postoperative Death

Supplement Figure 5. Clinician Predictions for Postoperative Acute Kidney Injury in Subgroup where Dichotomized ML Prediction Was Correct

Supplement Figure 6. Clinician Predictions for Postoperative Acute Kidney Injury in Subgroup where Dichotomized ML Prediction Was Incorrect

Supplement Table 3. Agreement Between Clinician and ML Predictions of Postoperative Acute Kidney Injury

Supplement Figure 7. Clinician Predictions for Postoperative Death Stratified by Clinician Reaction to ML

Supplement Figure 8. Clinician Predictions for Postoperative Acute Kidney Injury Stratified by Clinician Reaction to ML

Supplement Table 4. Effect of Clinician Reaction on Agreement Between Clinician and ML Predictions of Postoperative Death

Supplement Table 5. Effect of Clinician Reaction on Agreement Between Clinician and ML Predictions of Postoperative Acute Kidney Injury

Supplement Table 6. Predictive Performance of Clinician Predictions of Postoperative Death

Supplement Figure 9. Sensitivity and Subgroup Analyses for Prediction of Death

Supplement Table 7. Predictive Performance of Clinician Predictions of Postoperative Acute Kidney Injury

Supplement Figure 10. Sensitivity and Subgroup Analyses for Prediction of Acute Kidney Injury

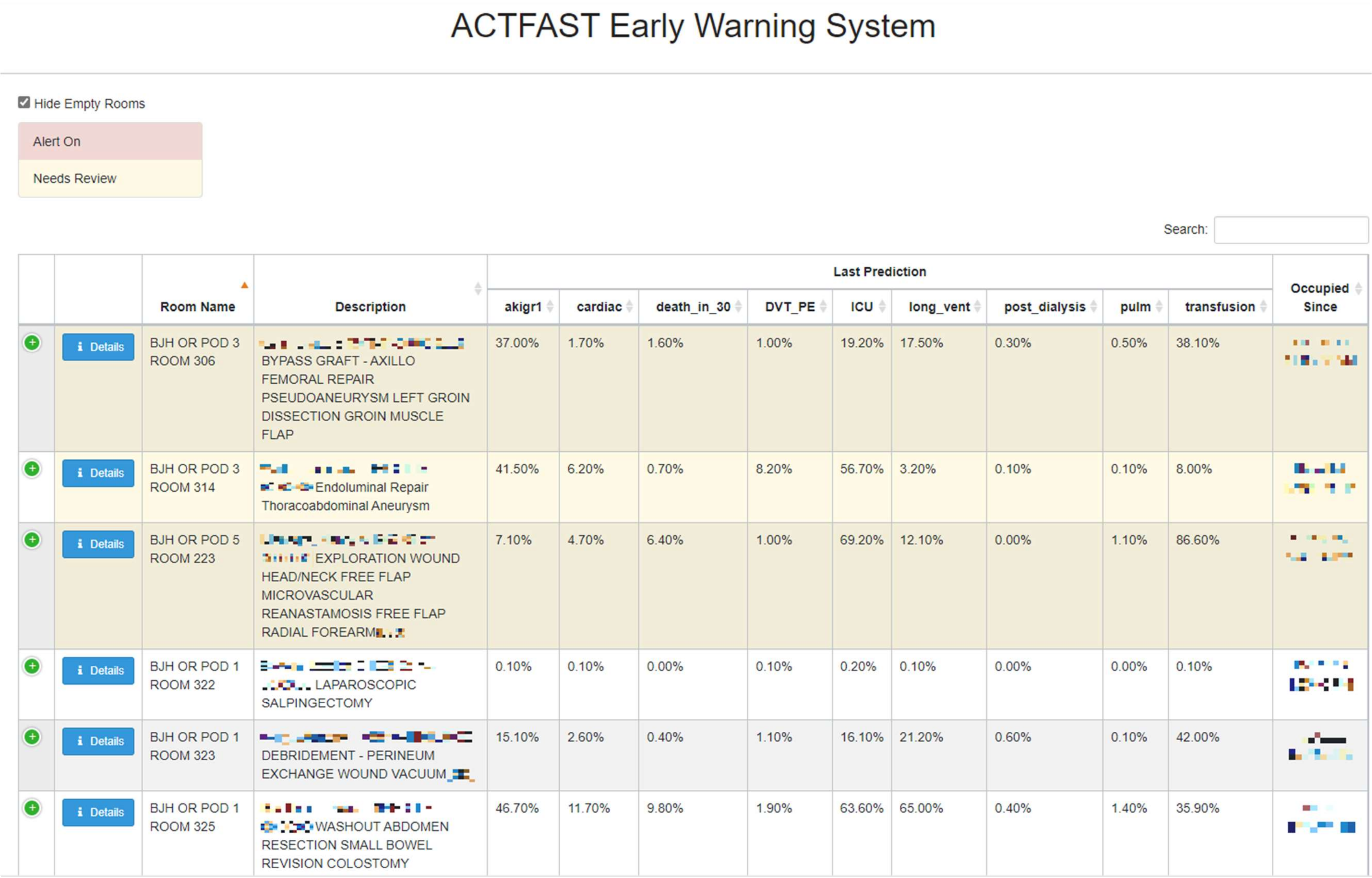

### ACTFAST Early Warning System

☑ Hide Empty Rooms

Alert On

Needs Review

Search:

| | | Room Name | Description | Last Prediction | | | | | | | | | Occupied Since |
|---|---|---|---|---|---|---|---|---|---|---|---|---|---|
| | | | | akigr1 | cardiac | death_in_30 | DVT_PE | ICU | long_vent | post_dialysis | pulm | transfusion | |
| + | Details | BJH OR POD 3 ROOM 306 | [illegible] BYPASS GRAFT - AXILLO FEMORAL REPAIR PSEUDOANEURYSM LEFT GROIN DISSECTION GROIN MUSCLE FLAP | 37.00% | 1.70% | 1.60% | 1.00% | 19.20% | 17.50% | 0.30% | 0.50% | 38.10% | [illegible] |
| + | Details | BJH OR POD 3 ROOM 314 | [illegible] Endoluminal Repair Thoracoabdominal Aneurysm | 41.50% | 6.20% | 0.70% | 8.20% | 56.70% | 3.20% | 0.10% | 0.10% | 8.00% | [illegible] |
| + | Details | BJH OR POD 5 ROOM 223 | [illegible] EXPLORATION WOUND HEAD/NECK FREE FLAP MICROVASCULAR REANASTAMOSIS FREE FLAP RADIAL FOREARM[illegible] | 7.10% | 4.70% | 6.40% | 1.00% | 69.20% | 12.10% | 0.00% | 1.10% | 86.60% | [illegible] |
| + | Details | BJH OR POD 1 ROOM 322 | [illegible] LAPAROSCOPIC SALPINGECTOMY | 0.10% | 0.10% | 0.00% | 0.10% | 0.20% | 0.10% | 0.00% | 0.00% | 0.10% | [illegible] |
| + | Details | BJH OR POD 1 ROOM 323 | [illegible] DEBRIDEMENT - PERINEUM EXCHANGE WOUND VACUUM[illegible] | 15.10% | 2.60% | 0.40% | 1.10% | 16.10% | 21.20% | 0.60% | 0.10% | 42.00% | [illegible] |
| + | Details | BJH OR POD 1 ROOM 325 | [illegible] WASHOUT ABDOMEN RESECTION SMALL BOWEL REVISION COLOSTOMY | 46.70% | 11.70% | 9.80% | 1.90% | 63.60% | 65.00% | 0.40% | 1.40% | 35.90% | [illegible] |

**Supplement Figure 1. Screenshot of User Interface.** Each row represents a patient, and each column is a different complication. Clicking on the “Details” button opens a view showing changes in predicted risk over time for a single patient and input features contributing to the predictions. Protected health information has been blurred.

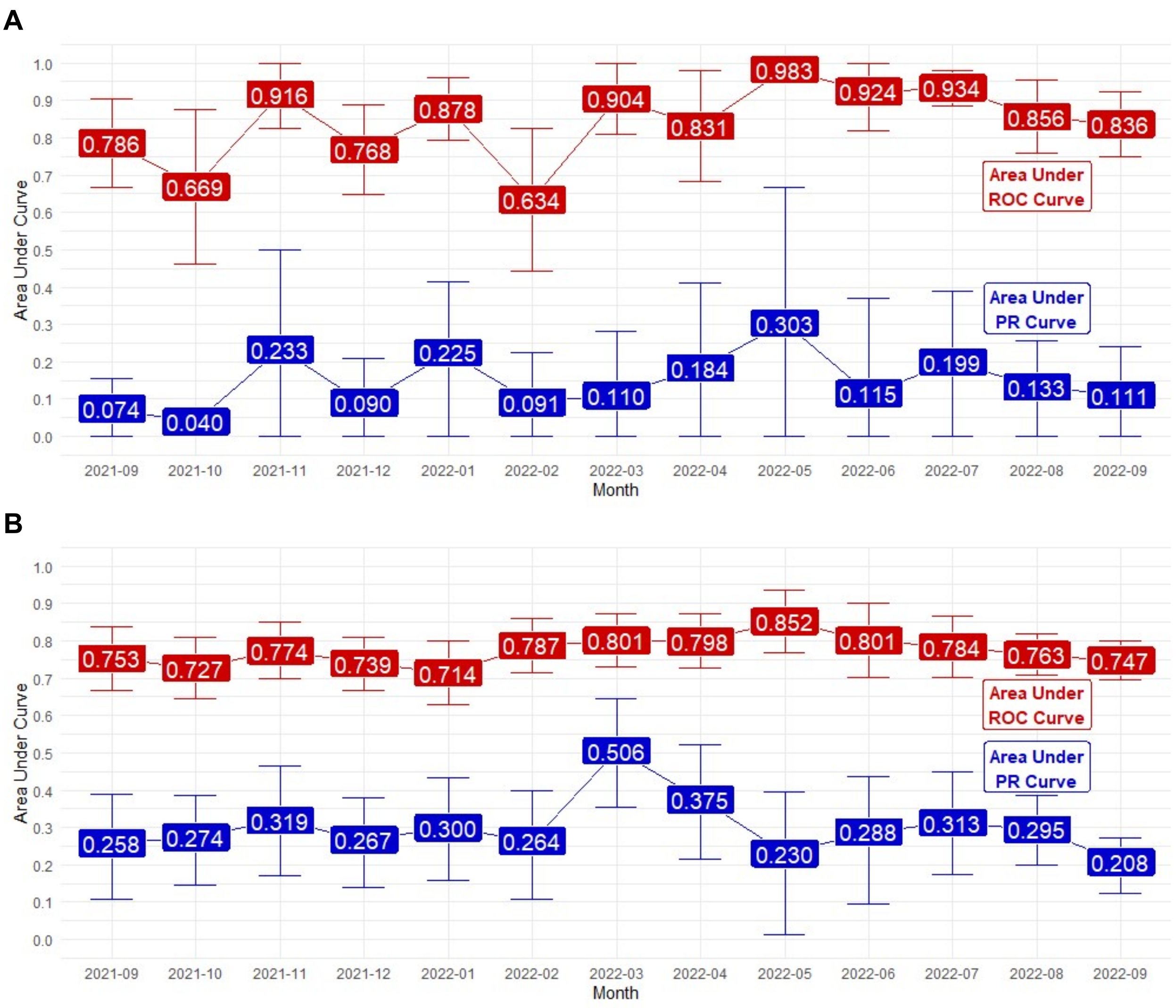


**Supplement Figure 2. Prospective Performance of Machine Learning Models Over Time.** (A) Performance of model predicting postoperative death. (B) Performance of model predicting postoperative acute kidney injury.

Abbreviations: ROC = receiver operating characteristic. PR = precision-recall.

**Supplement Table 1. Retrospective and Prospective Performance of Machine Learning Models**

| Outcome | Model Version | Performance Metric | Retrospective Performance | Prospective Performance |
|---|---|---|---|---|
| Death | Version 1 | AUROC | 0.94 | 0.81 |
| | | AUPRC | 0.16 | 0.12 |
| | Version 2 | AUROC | 0.91 | 0.84 |
| | | AUPRC | 0.27 | 0.13 |
| Acute Kidney Injury | Version 1 | AUROC | 0.80 | 0.74 |
| | | AUPRC | 0.28 | 0.27 |
| | Version 2 | AUROC | 0.90 | 0.78 |
| | | AUPRC | 0.66 | 0.29 |

AUROC = Area Under Receiver Operating Characteristic Curve.
AUPRC = Area Under Precision-Recall Curve

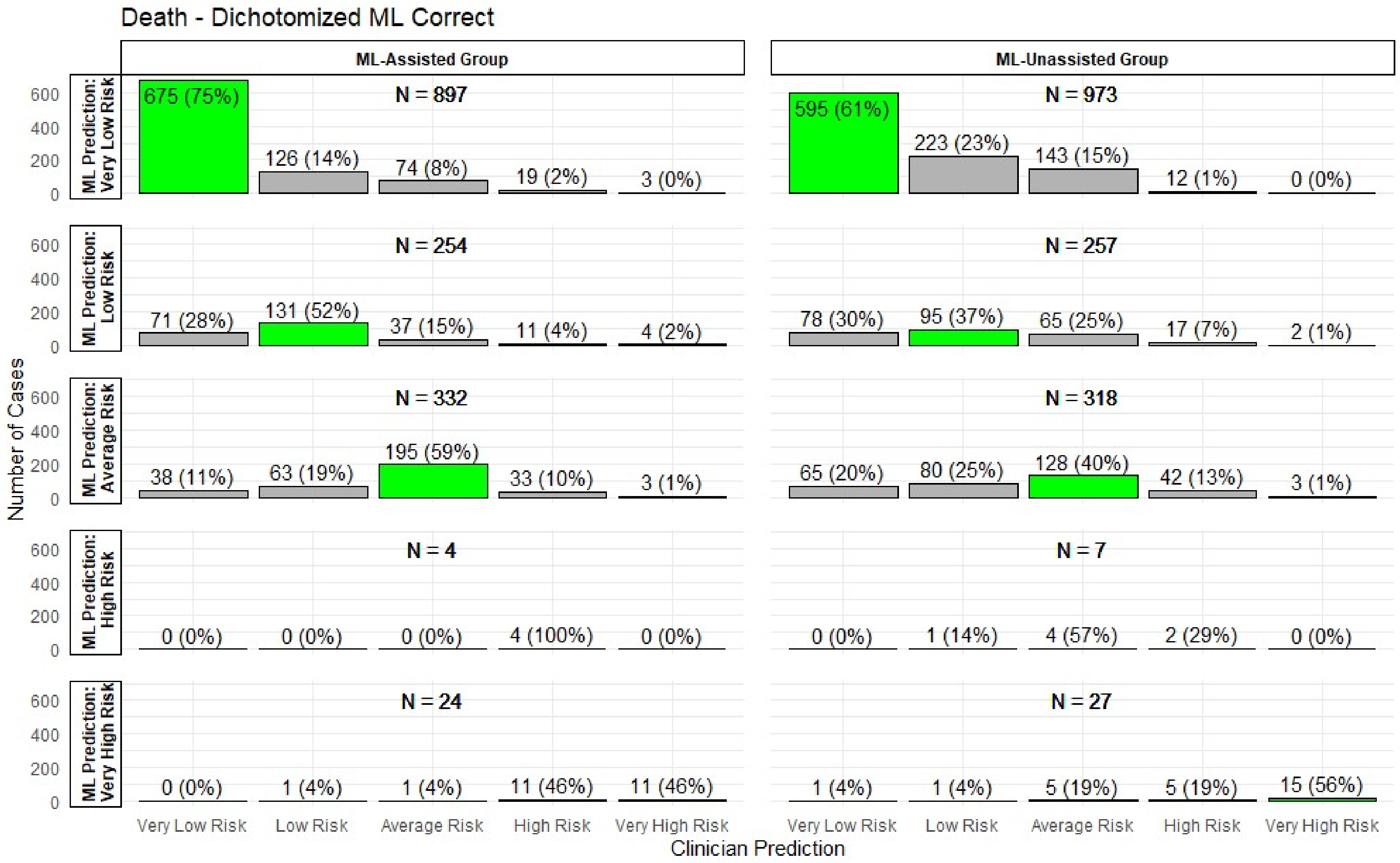


**Supplement Figure 3. Clinician Predictions for Postoperative Death in Subgroup where Dichotomized ML Prediction Was Correct.** ML predictions were dichotomized at the value that maximized the Youden index. Stratified by treatment allocation (ML-unassisted group versus ML-assisted group) and by ML prediction. Green bars represent cases where the clinician prediction matched the categorical ML prediction.

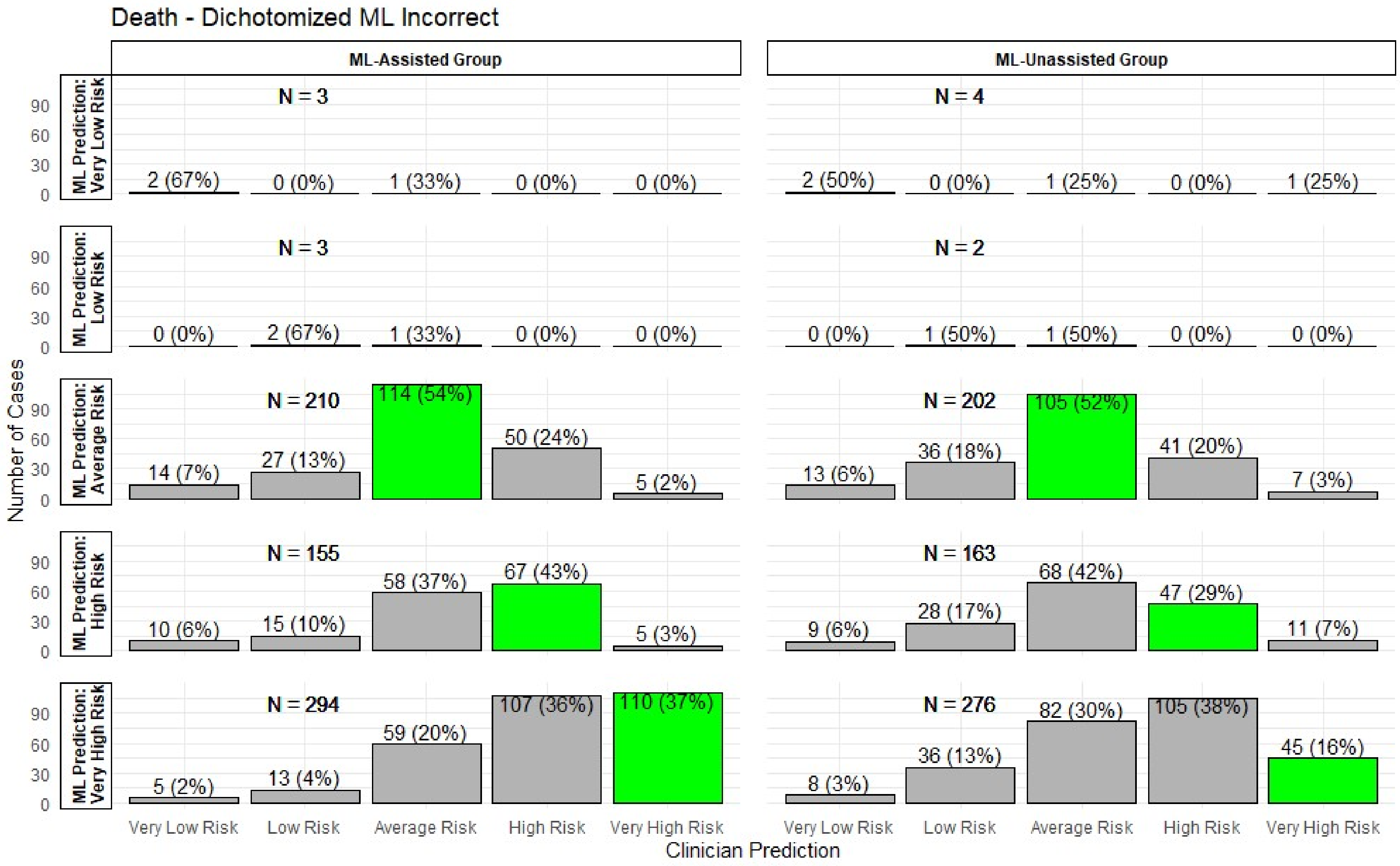


**Supplement Figure 4. Clinician Predictions for Postoperative Death in Subgroup where Dichotomized ML Prediction Was Incorrect.** ML predictions were dichotomized at the value that maximized the Youden index. Stratified by treatment allocation (ML-unassisted group versus ML-assisted group) and by ML prediction. Green bars represent cases where the clinician prediction matched the categorical ML prediction.

**Supplement Table 2. Agreement Between Clinician and ML Predictions of Postoperative Death**

| Cohort Name | Whole Cohort | Assisted Group | Unassisted Group | Difference between Groups |
|---|---|---|---|---|
| Entire Intention-to-Treat Cohort | | | | |
| N | 4459 | 2206 | 2253 | -- |
| Weighted Kappa [a] (95% CI) | 0.688 (0.687-0.688) | 0.751 (0.751-0.752) | 0.620 (0.619-0.620) | 0.13 (0.10 to 0.17) |
| Dichotomized ML Correct [b] | | | | |
| N | 3093 | 1511 | 1582 | -- |
| Weighted Kappa (95% CI) | 0.543 (0.542-0.544) | 0.615 (0.613-0.616) | 0.474 (0.473-0.476) | 0.14 (0.08 to 0.21) |
| Dichotomized ML Incorrect | | | | |
| N | 1312 | 665 | 647 | -- |
| Weighted Kappa (95% CI) | 0.274 (0.273-0.276) | 0.380 (0.378-0.382) | 0.177 (0.176-0.179) | 0.20 (0.12 to 0.28) |

[a] Weighted kappa measuring agreement, where the clinician prediction (5-point Likert scale) matched the categorical ML prediction (collapsed onto the same 5-point Likert scale). Kappa calculated using quadratic weights.
[b] Dichotomized at the value that maximized the Youden index.
Abbreviations: CI = Confidence interval. ML = Machine learning

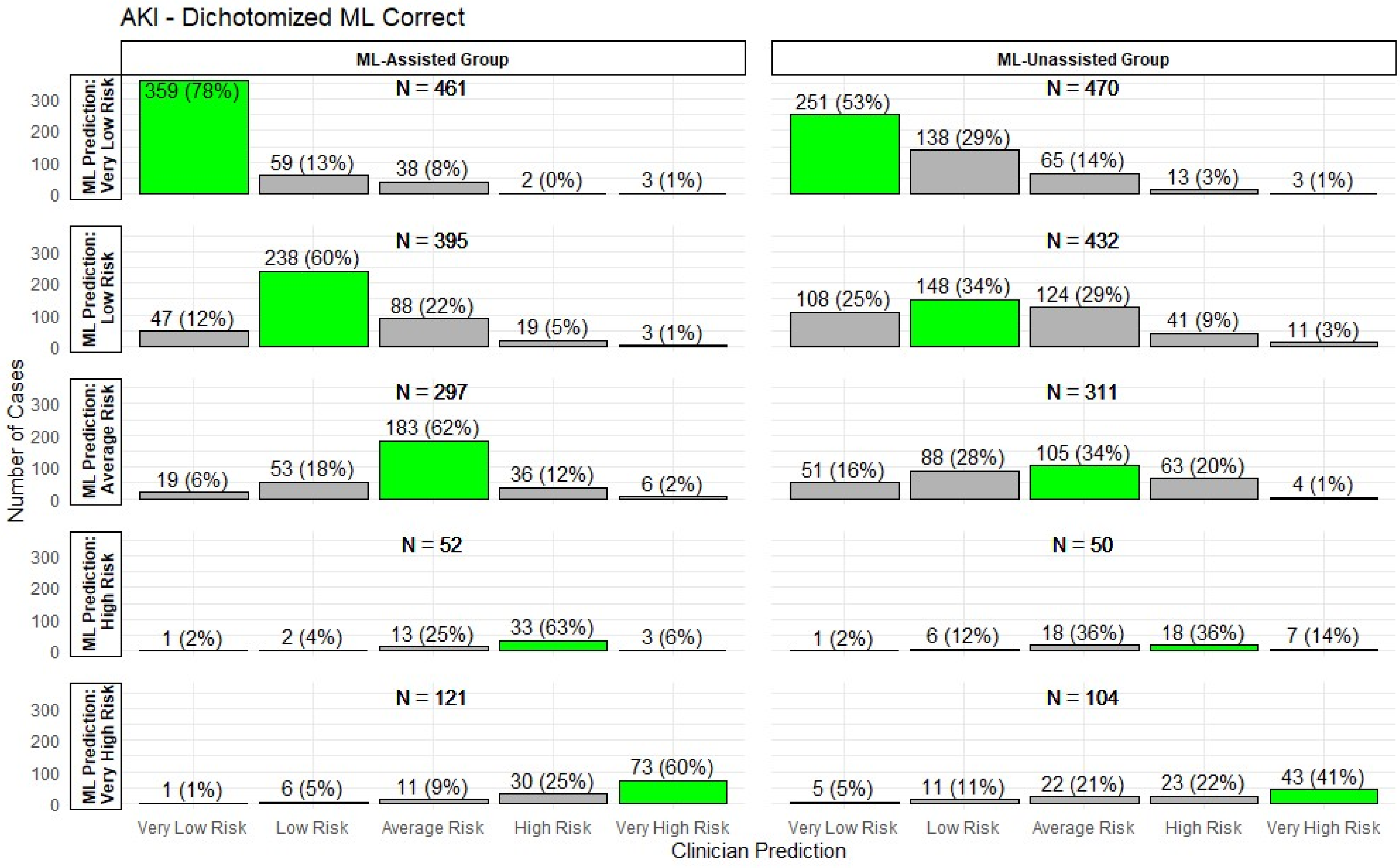


**Supplement Figure 5. Clinician Predictions for Postoperative Acute Kidney Injury in Subgroup where Dichotomized ML Prediction Was Correct.** ML predictions were dichotomized at the value that maximized the Youden index. Stratified by treatment allocation (ML-unassisted group versus ML-assisted group) and by ML prediction. Green bars represent cases where the clinician prediction matched the categorical ML prediction.

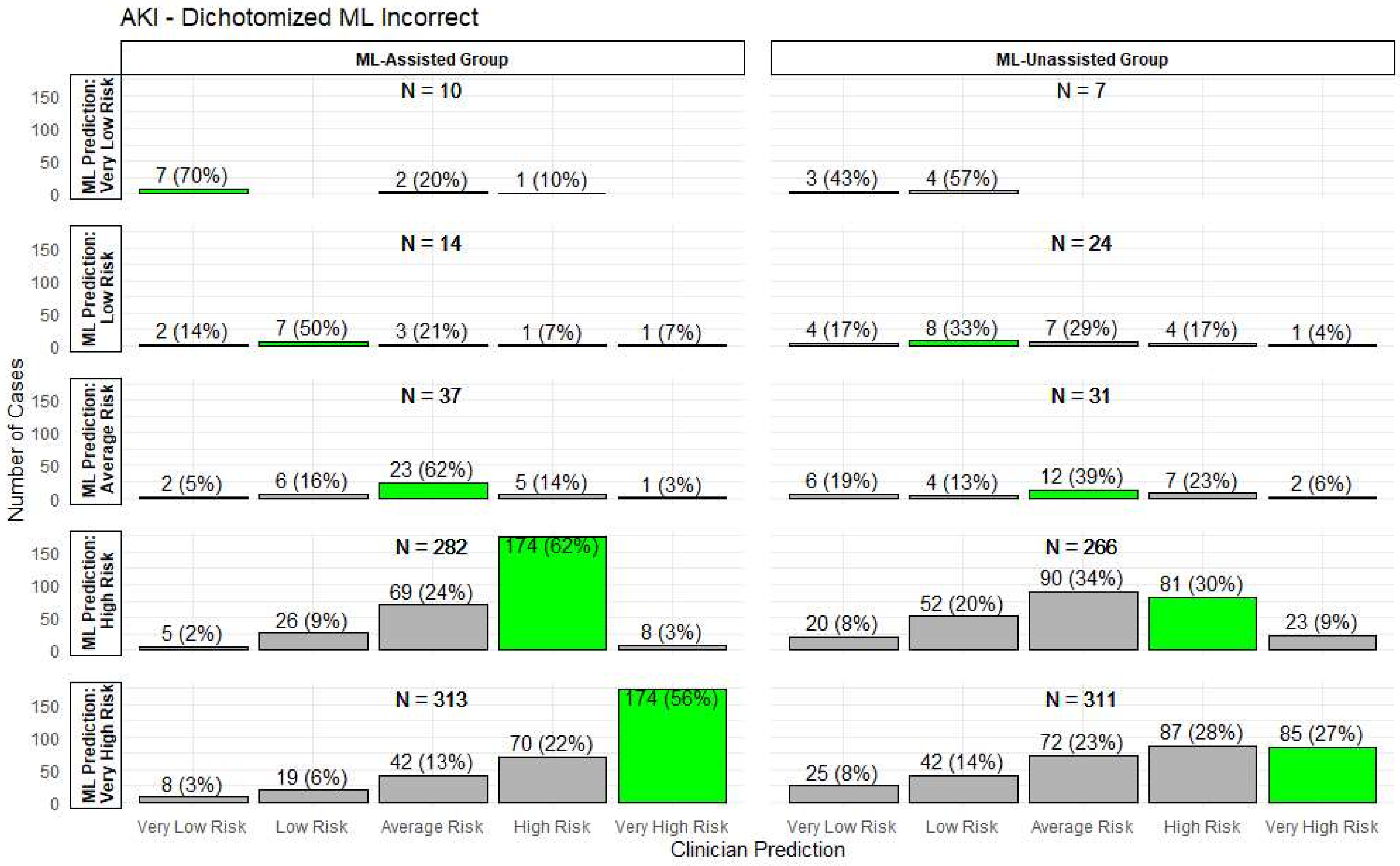


**Supplement Figure 6. Clinician Predictions for Postoperative Acute Kidney Injury in Subgroup where Dichotomized ML Prediction Was Incorrect.** ML predictions were dichotomized at the value that maximized the Youden index. Stratified by treatment allocation (ML-unassisted group versus ML-assisted group) and by ML prediction. Green bars represent cases where the clinician prediction matched the categorical ML prediction.

**Supplement Table 3. Agreement Between Clinician and ML Predictions of Postoperative Acute Kidney Injury**

| Cohort Name | Whole Cohort | Assisted Group | Unassisted Group | Difference between Groups |
|---|---|---|---|---|
| Entire Intention-to-Treat Cohort | | | | |
| N | 4055 | 2020 | 2035 | -- |
| Weighted Kappa [a] (95% CI) | 0.665 (0.665-0.666) | 0.787 (0.786-0.787) | 0.537 (0.536-0.538) | 0.25 (0.21 to 0.29) |
| Dichotomized ML Correct [b] | | | | |
| N | 2693 | 1326 | 1367 | -- |
| Weighted Kappa (95% CI) | 0.652 (0.652-0.653) | 0.774 (0.773-0.775) | 0.526 (0.525-0.527) | 0.25 (0.20 to 0.30) |
| Dichotomized ML Incorrect | | | | |
| N | 1295 | 656 | 639 | -- |
| Weighted Kappa (95% CI) | 0.253 (0.251-0.254) | 0.397 (0.395-0.400) | 0.157 (0.156-0.159) | 0.24 (0.15 to 0.33) |

[a] Weighted kappa measuring agreement, where the clinician prediction (5-point Likert scale) matched the categorical ML prediction (collapsed onto the same 5-point Likert scale). Kappa calculated using quadratic weights.
[b] Dichotomized at the value that maximized the Youden index.
Abbreviations: CI = Confidence interval. ML = Machine learning

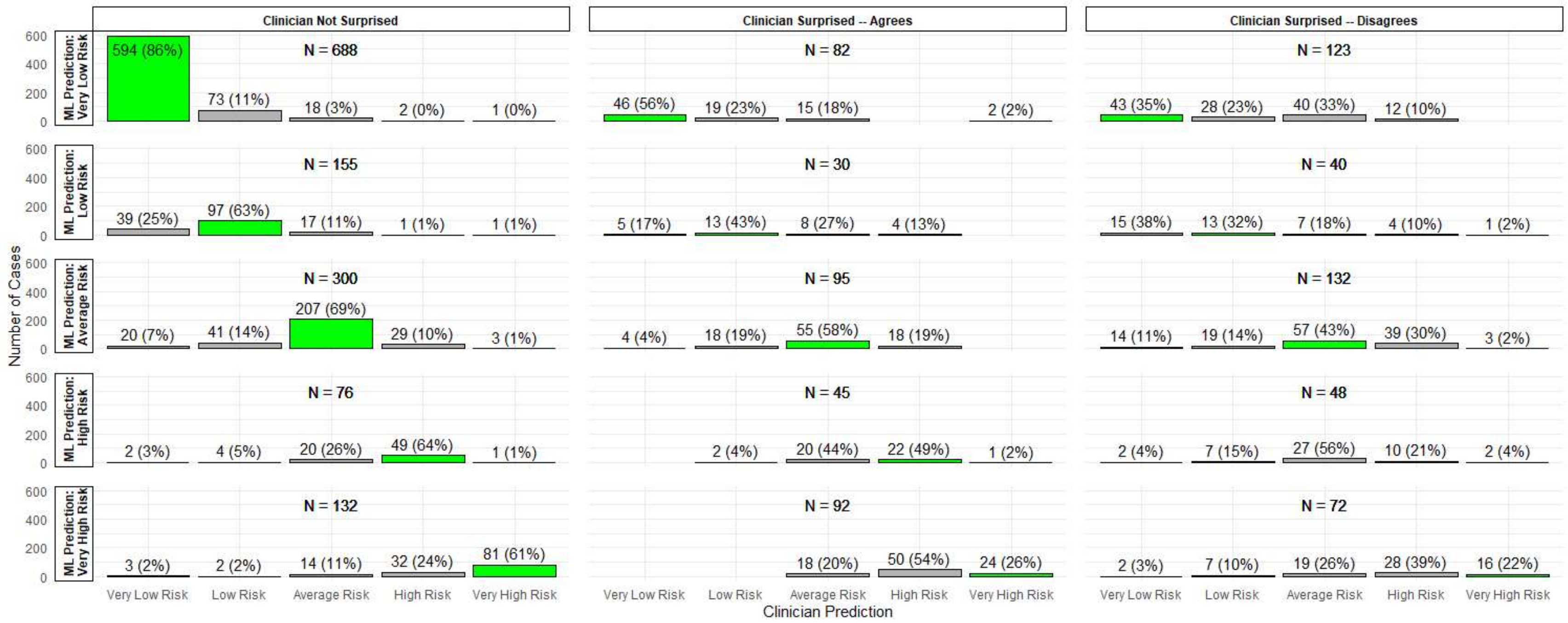


**Supplement Figure 7. Clinician Predictions for Postoperative Death Stratified by Clinician Reaction to ML.** Limited to cases where the clinician used ML and self-reported their reaction to it (not surprised, surprised and agrees, surprised and disagrees). Stratified by clinician reaction and by ML prediction. Green bars represent cases where the clinician prediction matched the categorical ML prediction.

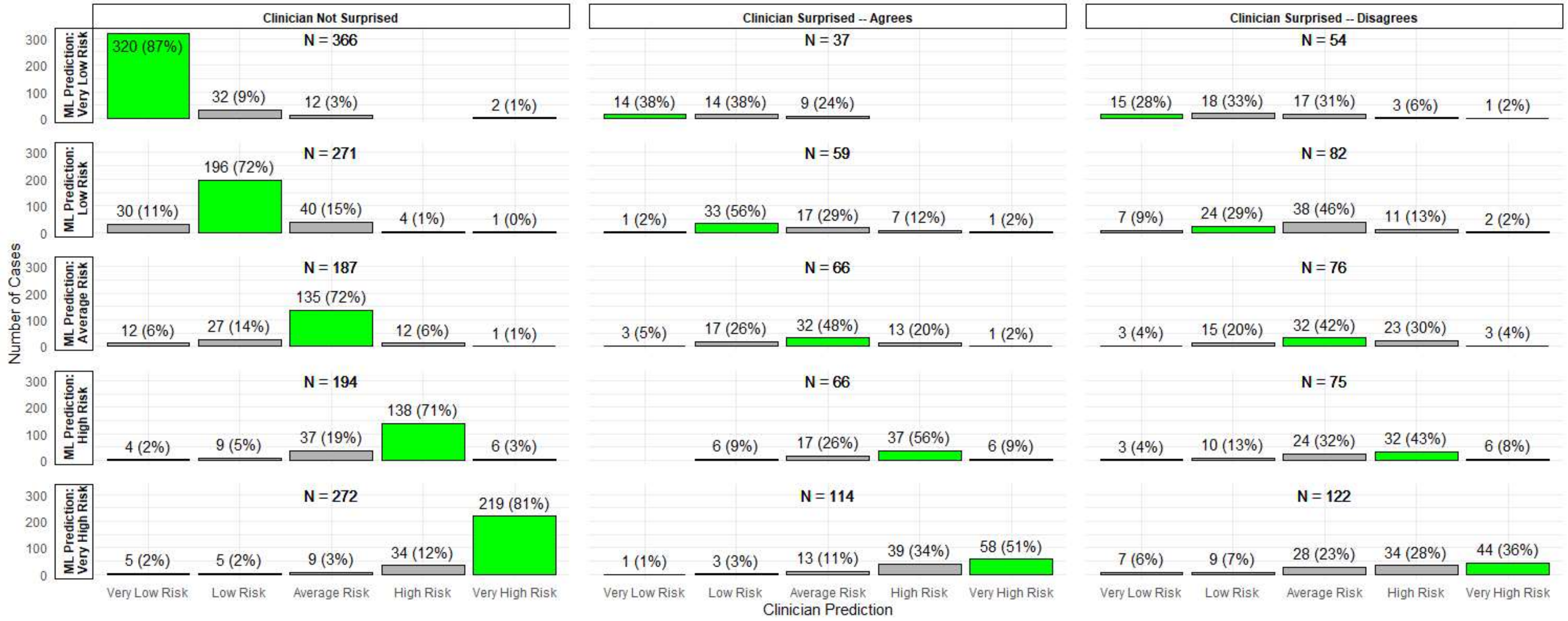


**Supplement Figure 8. Clinician Predictions for Postoperative Acute Kidney Injury Stratified by Clinician Reaction to ML.** Limited to cases where the clinician used ML and self-reported their reaction to it (not surprised, surprised and agrees, surprised and disagrees). Stratified by clinician reaction and by ML prediction. Green bars represent cases where the clinician prediction matched the categorical ML prediction.

**Supplement Table 4. Effect of Clinician Reaction on Agreement Between Clinician and ML Predictions of Postoperative Death**

| Clinician Reaction | N | Weighted Kappa [a] (95% CI) | Difference in Weighted Kappa [b] (95% CI) |
|---|---|---|---|
| Surprised by ML and Disagrees | 418 | 0.464 (0.462-0.467) | (Reference) |
| Surprised by ML but Agrees | 344 | 0.725 (0.723-0.727) | 0.26 (0.17 to 0.36) |
| Not Surprised by ML | 1363 | 0.866 (0.865-0.867) | 0.40 (0.33 to 0.48) |

[a] Weighted kappa measuring agreement, where the clinician prediction (5-point Likert scale) matched the categorical ML prediction (collapsed onto the same 5-point Likert scale). Kappa calculated using quadratic weights.
[b] Difference compared to the group where the clinician is surprised by ML and disagrees.
Abbreviations: CI = Confidence interval. ML = Machine learning

**Supplement Table 5. Effect of Clinician Reaction on Agreement Between Clinician and ML Predictions of Postoperative Acute Kidney Injury**

| Clinician Reaction | N | Weighted Kappa [a] (95% CI) | Difference in Weighted Kappa [b] (95% CI) |
|---|---|---|---|
| Surprised by ML and Disagrees | 415 | 0.460 (0.458-0.463) | (Reference) |
| Surprised by ML but Agrees | 345 | 0.706 (0.704-0.708) | 0.25 (0.15 to 0.35) |
| Not Surprised by ML | 1321 | 0.894 (0.893-0.894) | 0.43 (0.34 to 0.52) |

[a] Weighted kappa measuring agreement, where the clinician prediction (5-point Likert scale) matched the categorical ML prediction (collapsed onto the same 5-point Likert scale). Kappa calculated using quadratic weights.
[b] Difference compared to the group where the clinician is surprised by ML and disagrees.
Abbreviations: CI = Confidence interval. ML = Machine learning

**Supplement Table 6. Predictive Performance of Clinician Predictions of Postoperative Death**

| | ML-Assisted Group | | | | ML-Unassisted Group | | | |
|---|---|---|---|---|---|---|---|---|
| **Threshold [a]** | **Sensitivity** | **Specificity** | **PPV** | **NPV** | **Sensitivity** | **Specificity** | **PPV** | **NPV** |
| Very Low Risk | 46/46 (1.00) | 0/2160 (0.00) | 46/2206 (0.021) | 0/0 | 52/52 (1.00) | 0/2201 (0.00) | 52/2253 (0.023) | 0/0 |
| Low Risk | 44/46 (0.957) | 814/2160 (0.377) | 44/1390 (0.032) | 814/816 (0.998) | 49/52 (0.942) | 772/2201 (0.351) | 49/1478 (0.033) | 772/775 (0.996) |
| Average Risk | 40/46 (0.870) | 1194/2160 (0.553) | 40/1006 (0.040) | 1194/1200 (0.995) | 43/52 (0.827) | 1270/2201 (0.577) | 43/974 (0.044) | 1270/1279 (0.993) |
| High Risk | 31/46 (0.674) | 1741/2160 (0.806) | 31/450 (0.069) | 1741/1756 (0.991) | 27/52 (0.519) | 1867/2201 (0.848) | 27/361 (0.075) | 1867/1892 (0.987) |
| Very High Risk | 11/46 (0.239) | 2030/2160 (0.940) | 11/141 (0.078) | 2030/2065 (0.983) | 17/52 (0.327) | 2133/2201 (0.969) | 17/85 (0.200) | 2133/2168 (0.984) |

[a] Each row presents performance metrics when clinician predictions at or above the threshold category are treated as positive predictions.
Abbreviations: ML = Machine learning. PPV = Positive predictive value. NPV = Negative predictive value.

| Analysis | N | ML-Assisted Group AUROC (95% CI) | ML-Unassisted Group AUROC (95% CI) |
|---|---|---|---|
| Intention to Treat | 4459 | 0.793 (0.735-0.851) | 0.780 (0.717-0.842) |
| Per Protocol | 3587 | 0.795 (0.723-0.868) | 0.785 (0.721-0.849) |
| As Treated | 4230 | 0.798 (0.740-0.857) | 0.768 (0.703-0.833) |
| Models Version 1 | 1605 | 0.783 (0.716-0.849) | 0.820 (0.735-0.905) |
| Models Version 2 | 2854 | 0.819 (0.744-0.895) | 0.781 (0.701-0.862) |
| Female | 2339 | 0.813 (0.739-0.887) | 0.759 (0.654-0.864) |
| Male | 2120 | 0.774 (0.688-0.860) | 0.787 (0.711-0.864) |
| White Race | 3402 | 0.799 (0.736-0.863) | 0.778 (0.702-0.854) |
| Black Race | 912 | 0.770 (0.639-0.902) | 0.776 (0.640-0.911) |
| TECTONICS Telemedicine Group | 2229 | 0.819 (0.752-0.887) | 0.746 (0.646-0.845) |
| TECTONICS Usual Care Group | 2230 | 0.765 (0.669-0.861) | 0.817 (0.745-0.889) |
| MD/DO - Attending | 1416 | 0.810 (0.681-0.940) | 0.829 (0.757-0.901) |
| MD/DO - CA3 Resident | 1068 | 0.718 (0.619-0.818) | 0.790 (0.560-1.000) |
| MD/DO - Intern Level | 1246 | 0.841 (0.707-0.974) | 0.854 (0.769-0.938) |
| CRNA | 486 | 0.818 (0.741-0.896) | 0.835 (0.580-1.000) |
| SRNA | 243 | 0.867 (NA-NA) | 0.635 (0.333-0.937) |
| Correct ML Prediction | 3093 | 0.957 (0.933-0.981) | 0.884 (0.835-0.932) |
| Incorrect ML Prediction | 1312 | 0.215 (0.093-0.337) | 0.410 (0.152-0.668) |

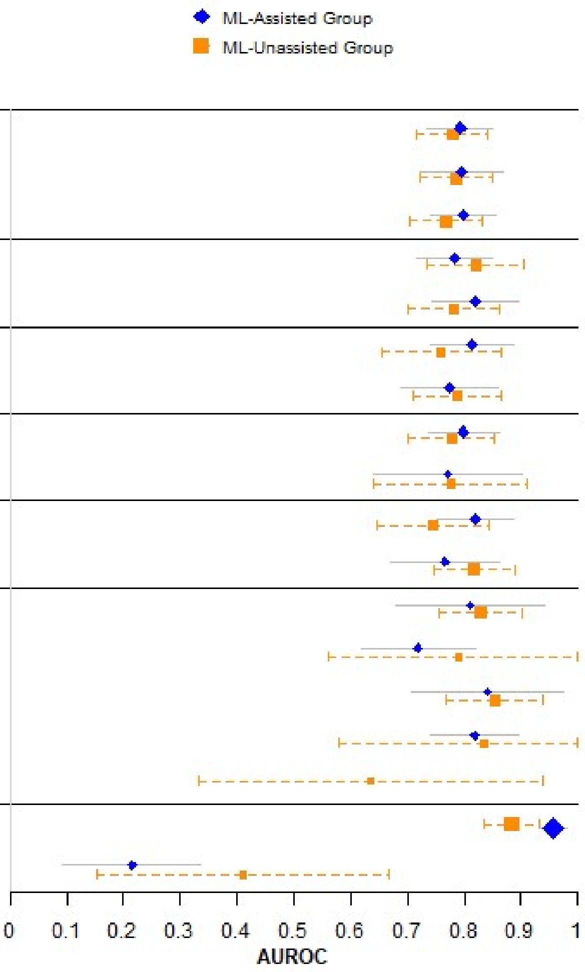


**Supplement Figure 9. Sensitivity and Subgroup Analyses for Prediction of Death**
Abbreviations: ML = Machine learning. AUROC = Area under receiver operating characteristic curve. CRNA = certified registered nurse anesthetist. SRNA = student registered nurse anesthetist.

**Supplement Table 7. Predictive Performance of Clinician Predictions of Postoperative Acute Kidney Injury**

| | ML-Assisted Group | | | | ML-Unassisted Group | | | |
|---|---|---|---|---|---|---|---|---|
| **Threshold [a]** | **Sensitivity** | **Specificity** | **PPV** | **NPV** | **Sensitivity** | **Specificity** | **PPV** | **NPV** |
| Very Low Risk | 234/234 (1.00) | 0/1786 (0.00) | 234/2020 (0.116) | 0/0 (NaN) | 216/216 (1.00) | 0/1819 (0.00) | 216/2035 (0.106) | 0/0 (NaN) |
| Low Risk | 222/234 (0.949) | 441/1786 (0.247) | 222/1567 (0.142) | 441/453 (0.974) | 197/216 (0.912) | 458/1819 (0.252) | 197/1558 (0.126) | 458/477 (0.960) |
| Average Risk | 203/234 (0.868) | 847/1786 (0.474) | 203/1142 (0.178) | 847/878 (0.965) | 165/216 (0.764) | 930/1819 (0.511) | 165/1054 (0.157) | 930/981 (0.948) |
| High Risk | 151/234 (0.645) | 1284/1786 (0.719) | 151/653 (0.231) | 1284/1367 (0.939) | 107/216 (0.495) | 1395/1819 (0.767) | 107/531 (0.202) | 1395/1504 (0.928) |
| Very High Risk | 78/234 (0.333) | 1590/1786 (0.890) | 78/274 (0.285) | 1590/1746 (0.911) | 54/216 (0.250) | 1690/1819 (0.929) | 54/183 (0.295) | 1690/1852 (0.913) |

[a] Each row presents performance metrics when clinician predictions at or above the threshold category are treated as positive predictions.

Abbreviations: ML = Machine learning. PPV = Positive predictive value. NPV = Negative predictive value.

| Analysis | N | ML-Assisted Group AUROC (95% CI) | ML-Unassisted Group AUROC (95% CI) |
|---|---|---|---|
| Intention to Treat | 4055 | 0.734 (0.703-0.766) | 0.688 (0.652-0.725) |
| Per Protocol | 3269 | 0.733 (0.699-0.766) | 0.693 (0.653-0.733) |
| As Treated | 3844 | 0.731 (0.699-0.763) | 0.694 (0.657-0.732) |
| Exclude If No Postoperative Creatinine | 2960 | 0.676 (0.640-0.711) | 0.644 (0.605-0.684) |
| Models Version 1 | 1443 | 0.733 (0.682-0.785) | 0.665 (0.600-0.730) |
| Models Version 2 | 2612 | 0.736 (0.696-0.775) | 0.707 (0.664-0.751) |
| Female | 2179 | 0.754 (0.704-0.803) | 0.705 (0.651-0.759) |
| Male | 1876 | 0.705 (0.663-0.748) | 0.665 (0.614-0.716) |
| White Race | 3171 | 0.732 (0.697-0.768) | 0.691 (0.647-0.735) |
| Black Race | 750 | 0.746 (0.673-0.819) | 0.685 (0.612-0.757) |
| TECTONICS Telemedicine Group | 2017 | 0.718 (0.672-0.764) | 0.696 (0.638-0.755) |
| TECTONICS Usual Care Group | 2038 | 0.750 (0.707-0.792) | 0.681 (0.634-0.728) |
| MD/DO - Attending | 1277 | 0.735 (0.679-0.790) | 0.674 (0.608-0.741) |
| MD/DO - CA3 Resident | 955 | 0.709 (0.633-0.784) | 0.649 (0.569-0.730) |
| MD/DO - Intern Level | 1155 | 0.748 (0.699-0.797) | 0.741 (0.688-0.794) |
| CRNA | 448 | 0.736 (0.627-0.844) | 0.725 (0.562-0.887) |
| SRNA | 220 | 0.659 (0.424-0.894) | 0.673 (0.530-0.817) |
| Correct ML Prediction | 2693 | 0.926 (0.905-0.947) | 0.832 (0.798-0.866) |
| Incorrect ML Prediction | 1295 | 0.198 (0.143-0.253) | 0.330 (0.261-0.399) |

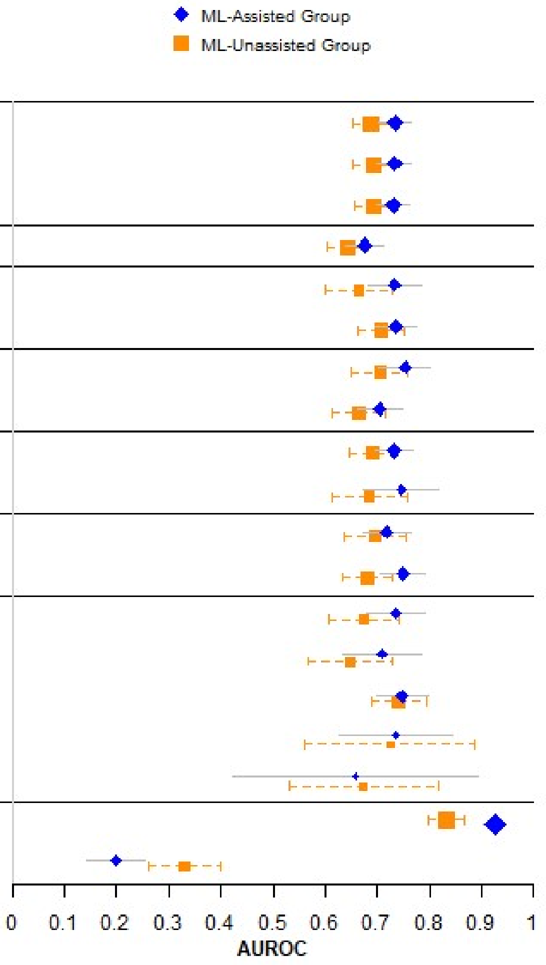


**Supplement Figure 10. Sensitivity and Subgroup Analyses for Prediction of Acute Kidney Injury**
Abbreviations: ML = Machine learning. AUROC = Area under receiver operating characteristic curve. CRNA = certified registered nurse anesthetist. SRNA = student registered nurse anesthetist
