## Supplementary material for "Effect of Machine Learning on Anaesthesiology Clinician Prediction of Postoperative Complications: The Perioperative ORACLE Randomised Clinical Trial": CONSORT-AI Checklist

Checklist of information to include when reporting randomised trials of AI interventions

| Section | Item | CONSORT 2010 Item^a^ | CONSORT-AI Item | | Addressed on Page No^b^ |
| --- | --- | --- | --- | --- | --- |
| Title and Abstract | | | | | |
| **Title and Abstract** | 1a | Identification as a randomised trial in the title | CONSORT-AI 1a,b Elaboration | (i) Indicate that the intervention involves artificial intelligence/machine learning in the title and/or abstract and specify the type of model. | 1 |
|  | 1b | Structured summary of trial design, methods, results, and conclusions (for specific guidance see CONSORT for abstracts) |  | (ii) State the intended use of the AI intervention within the trial in the title and/or abstract. | 2 |
| Introduction | | | | | |
| **Background and objectives** | 2a | Scientific background and explanation of rationale | CONSORT-AI 2a (i) Extension | Explain the intended use of the AI intervention in the context of the clinical pathway, including its purpose and its intended users (e.g. healthcare professionals, patients, public). | 3 |
|  | 2b | Specific objectives or hypotheses |  |  | 3 |
| Methods | | | | | |
| **Trial design** | 3a | Description of trial design (such as parallel, factorial) including allocation ratio |  |  | 4 |
|  | 3b | Important changes to methods after trial commencement (such as eligibility criteria), with reasons |  |  | NA |
| **Participants** | 4a | Eligibility criteria for participants | CONSORT-AI 4a (i) Elaboration | State the inclusion and exclusion criteria at the level of participants. | 4-5 |
|  |  |  | CONSORT-AI 4a (ii) Extension | State the inclusion and exclusion criteria at the level of the input data. | 5 |
|  | 4b | Settings and locations where the data were collected | CONSORT-AI 4b Extension | Describe how the AI intervention was integrated into the trial setting, including any onsite or offsite requirements. | 5 |
| **Interventions** | 5 | The interventions for each group with sufficient details to allow replication, including how and when they were actually administered | CONSORT-AI 5 (i) Extension | State which version of the AI algorithm was used. | 5 |
|  |  |  | CONSORT-AI 5 (ii) Extension | Describe how the input data were acquired and selected for the AI intervention. | 5 |
|  |  |  | CONSORT-AI 5 (iii) Extension | Describe how poor quality or unavailable input data were assessed and handled. | 5 |
|  |  |  | CONSORT-AI 5 (iv) Extension. | Specify whether there was human-AI interaction in the handling of the input data, and what level of expertise was required of users. | 6 |
|  |  |  | CONSORT-AI 5 (v) Extension | Specify the output of the AI intervention | 5 |
|  |  |  | CONSORT-AI 5 (vi) Extension | Explain how the AI intervention’s outputs contributed to decision-making or other elements of clinical practice. | 6 |
| **Outcomes** | 6a | Completely defined pre-specified primary and secondary outcome measures, including how and when they were assessed |  |  | 6-7 |
|  | 6b | Any changes to trial outcomes after the trial commenced, with reasons |  |  | NA |
| **Sample size** | 7a | How sample size was determined |  |  | 7 |
|  | 7b | When applicable, explanation of any interim analyses and stopping guidelines |  |  | NA |
| Randomisation | | | | | |
| **Sequence generation** | 8a | Method used to generate the random allocation sequence |  |  | 6 |
|  | 8b | Type of randomisation; details of any restriction (such as blocking and block size) |  |  | 6 |
| **Allocation concealment mechanism** | 9 | Mechanism used to implement the random allocation sequence (such as sequentially numbered containers), describing any steps taken to conceal the sequence until interventions were assigned |  |  | 6 |
| **Implementation** | 10 | Who generated the random allocation sequence, who enrolled participants, and who assigned participants to interventions |  |  | 6 |
| **Blinding** | 11a | If done, who was blinded after assignment to interventions (for example, participants, care providers, those assessing outcomes) and how |  |  | 6 |
|  | 11b | If relevant, description of the similarity of interventions |  |  | 6 |
| **Statistical methods** | 12a | Statistical methods used to compare groups for primary and secondary outcomes |  |  | 7 |
|  | 12b | Methods for additional analyses, such as subgroup analyses and adjusted analyses |  |  | 7-8 |
| Results | | | | | |
| **Participant flow** (a diagram is strongly recommended) | 13a | For each group, the numbers of participants who were randomly assigned, received intended treatment, and were analysed for the primary outcome |  |  | Fig 1 |
|  | 13b | For each group, losses and exclusions after randomisation, together with reasons |  |  | Fig 1 |
| **Recruitment** | 14a | Dates defining the periods of recruitment and follow-up |  |  | 9 |
|  | 14b | Why the trial ended or was stopped |  |  | 9 |
| **Baseline data** | 15 | A table showing baseline demographic and clinical characteristics for each group |  |  | Table 1 |
| **Numbers analysed** | 16 | For each group, number of participants (denominator) included in each analysis and whether the analysis was by original assigned groups |  |  | 9-10, Fig 1 |
| **Outcomes and estimation** | 17a | For each primary and secondary outcome, results for each group, and the estimated effect size and its precision (such as 95% confidence interval) |  |  | 9-10 |
|  | 17b | For binary outcomes, presentation of both absolute and relative effect sizes is recommended |  |  | 9-10 |
| **Ancillary analyses** | 18 | Results of any other analyses performed, including subgroup analyses and adjusted analyses, distinguishing pre-specified from exploratory |  |  | 10, Supp Figs 9-10 |
| **Harms** | 19 | All important harms or unintended effects in each group (for specific guidance see CONSORT for harms) | CONSORT-AI 19 Extension | Describe results of any analysis of performance errors and how errors were identified, where applicable. If no such analysis was planned or done, explain why not. | 10-11, Supp Figs 3-6 |
| Discussion | | | | | |
| **Limitations** | 20 | Trial limitations, addressing sources of potential bias, imprecision, and, if relevant, multiplicity of analyses |  |  | 13 |
| **Generalisability** | 21 | Generalisability (external validity, applicability) of the trial findings |  |  | 13 |
| **Interpretation** | 22 | Interpretation consistent with results, balancing benefits and harms, and considering other relevant evidence |  |  | 13-14 |
| Other Information | | | | | |
| **Registration** | 23 | Registration number and name of trial registry |  |  | 2 |
| **Protocol** | 24 | Where the full trial protocol can be accessed, if available |  |  | 4 |
| **Funding** | 25 | Sources of funding and other support (such as supply of drugs), role of funders | CONSORT-AI 25 Extension. | State whether and how the AI intervention and/or its code can be accessed, including any restrictions to access or re-use. | 15 |

^a^ We strongly recommend reading this statement in conjunction with the CONSORT 2010 Explanation and Elaboration for important clarifications on all the items.

^b^ Indicates page numbers to be completed by authors during protocol development.
